# Perceptions of non-invasive brain stimulation and barriers to it’s clinical translation: a multi-stakeholder focus group study

**DOI:** 10.64898/2026.08.24.26361180

**Authors:** Matthew Weightman, Barbara Robinson, Hanna Smyth, Anton Pick, Elly Martin, Jessica Walsh, Charlotte J Stagg, Melanie K Fleming

## Abstract

**Objectives:** Non-invasive brain stimulation (NIBS) holds significant promise for treating neurological and neuropsychiatric conditions, yet translation into routine clinical practice remains limited. We aimed to explore stakeholder perceptions of NIBS and barriers to its clinical adoption.

**Methods:** We conducted focus-group interviews with 33 participants across three key stakeholder groups in the UK: (1) people with lived experience of brain injury, depression, or dementia; (2) healthcare professionals; and (3) researchers. Reflexive thematic analysis was used to identify themes in the data.

**Findings:** Seven key themes emerged spanning preferences, hope and disappointment, communication, accessibility, infrastructure, ethical/regulatory uncertainty, and the evidence base. Across groups, NIBS was viewed positively and with cautious optimism, but substantial barriers were highlighted, including limited public and clinical awareness, challenges in demonstrating cost-effectiveness, infrastructure constraints, and difficulties navigating regulatory and translational pathways. Participants emphasised the importance of clear communication, improved education, and stronger interdisciplinary collaboration to support adoption. Notably, stakeholders prioritised evidence of clinical efficacy and usability over detailed mechanistic understanding.

**Conclusions:** These findings provide actionable insights into the translational gap in NIBS and highlight priorities for facilitating its integration into clinical care.

## Introduction

With an ageing society, neurological, neurodegenerative, and neuropsychiatric conditions are an increasing global burden, carrying immense personal and societal costs ^1^. Treatments to alleviate symptoms and improve function are therefore urgently required. Non-invasive brain stimulation (NIBS) offers a promising, scalable way to directly and precisely interact with brain activity to treat the symptoms and alter the course of neurological and psychiatric diseases. Unlike pharmacological approaches, NIBS can target specific brain regions to modulate ongoing network dynamics with exceptional spatial and temporal precision, opening the door to more effective and personalised interventions ^2^.

However, fulfilling this promise requires not only substantial technical development, but also the successful translation of research into clinical practice. Despite decades of research and technological advances, very few NIBS treatments currently exist in clinical care across the world. There are significant barriers to scaling-up brain stimulation device use, including limited understanding of efficacy and potential for personalisation; side effects; and accessibility considerations ^3–6^. Indeed, perceptions may also differ across stakeholders based on differences in beliefs, concerns, and knowledge. It is therefore vital to understand perceptions of different types of NIBS interventions, for people with experience of different neurological, neurodegenerative and neuropsychiatric conditions as well as from those responsible for developing and delivering treatments now and in the future.

A systematic review of NIBS for therapeutic use ^7^ reported that most patients considered repetitive Transcranial Magnetic Stimulation (TMS) to be safe for psychiatric treatment but had limited understanding of its efficacy. A follow-up study of TMS for chronic pain ^8^ identified several key themes influencing acceptance. These included perceptions of the equipment and familiarity with technology, accessibility, understanding of their condition and the treatment rationale. Similarly, a recent survey found that patients and the public had a low baseline knowledge of NIBS techniques, but that even brief information reduced confusion, and increased their excitement, optimism and comfort ^9^. There was some indication for a preference of transcranial ultrasound stimulation (TUS) over magnetic or electrical alternatives, although the underlying reasons remain unclear.

Building on these findings, we aimed to gain a deeper understanding of perceptions of NIBS technologies and the barriers to clinical translation across a range of stakeholders in the United Kingdom. By synthesizing perspectives from a range of stakeholders, including patients, clinicians, and researchers we aimed to identify system level translational barriers not visible in single stakeholder studies. We focus on NIBS techniques including transcranial magnetic (TMS), electrical (tES) and ultrasound (TUS) stimulation. To understand perceptions from across a range of neurological, neurodegenerative, and neuropsychiatric conditions we interviewed people with lived experience of acquired brain injury, depression, and dementia. We selected these three conditions for their high prevalence and diversity of lived experience. To supplement the patient/public point-of-view, we also aimed to gather insights from healthcare professionals and NIBS researchers.

## Methods

### Participants

A convenience sample of participants was recruited through our research databases, as well as through informal networks, charities, conferences and online advertising. Inclusion criteria were being aged 18 years or over, able/willing to provide informed consent for participation in a focus group interview and living in the United Kingdom.

For the lived experience (LE) group, participants needed to have lived experience of neurological, psychiatric or neurodegenerative brain conditions, in particular acquired brain injury (including stroke, trauma and brain bleeds), depression (or low mood), and dementia. This included people with direct experience (themselves) or relatives/friends who have experience caring for someone with any of these conditions.

For the healthcare professional (HCP) group, participants needed to have experience working with people who have neurological, neuropsychiatric or neurodegenerative conditions (in particular brain injury, depression or dementia). We deliberately sought to include people from a range of clinical roles, including medical and allied health.

Finally, for the researcher (R) group, participants needed to have experience conducting research related to non-invasive brain stimulation techniques, particularly TMS, TUS, tES and/or neurological, neuropsychiatric or neurodegenerative conditions (including brain injury, depression or dementia). We deliberately sought to include people from a range of research backgrounds, including neuroscience, physics, engineering, law.

In total, 38 participants provided informed consent. Of these, five failed to attend the session (2 no reason given, 1 logistical difficulties, 2 personal reasons) and one was unable to be scheduled due to restrictions on availability. We included options for both in person and online interviews based on preference and to ensure that travel requirements were not a barrier to participation. The breakdown of focus group sizes and modes of delivery (in person / online) are in table 1. Where possible the groupings were kept separate (i.e. LE participants were not included in HCP or R focus groups) to ensure openness of conversations (e.g. LE participants might not be willing to make certain comments if a HCP was in the same interview) and to avoid issues with group-specific jargon (e.g. R group participants might use scientific language that could be difficult for LE participants to follow). However, some participants identified as fitting more than one group and were thus scheduled for the interview grouping that fitted best with their availability.

**Table 1.**
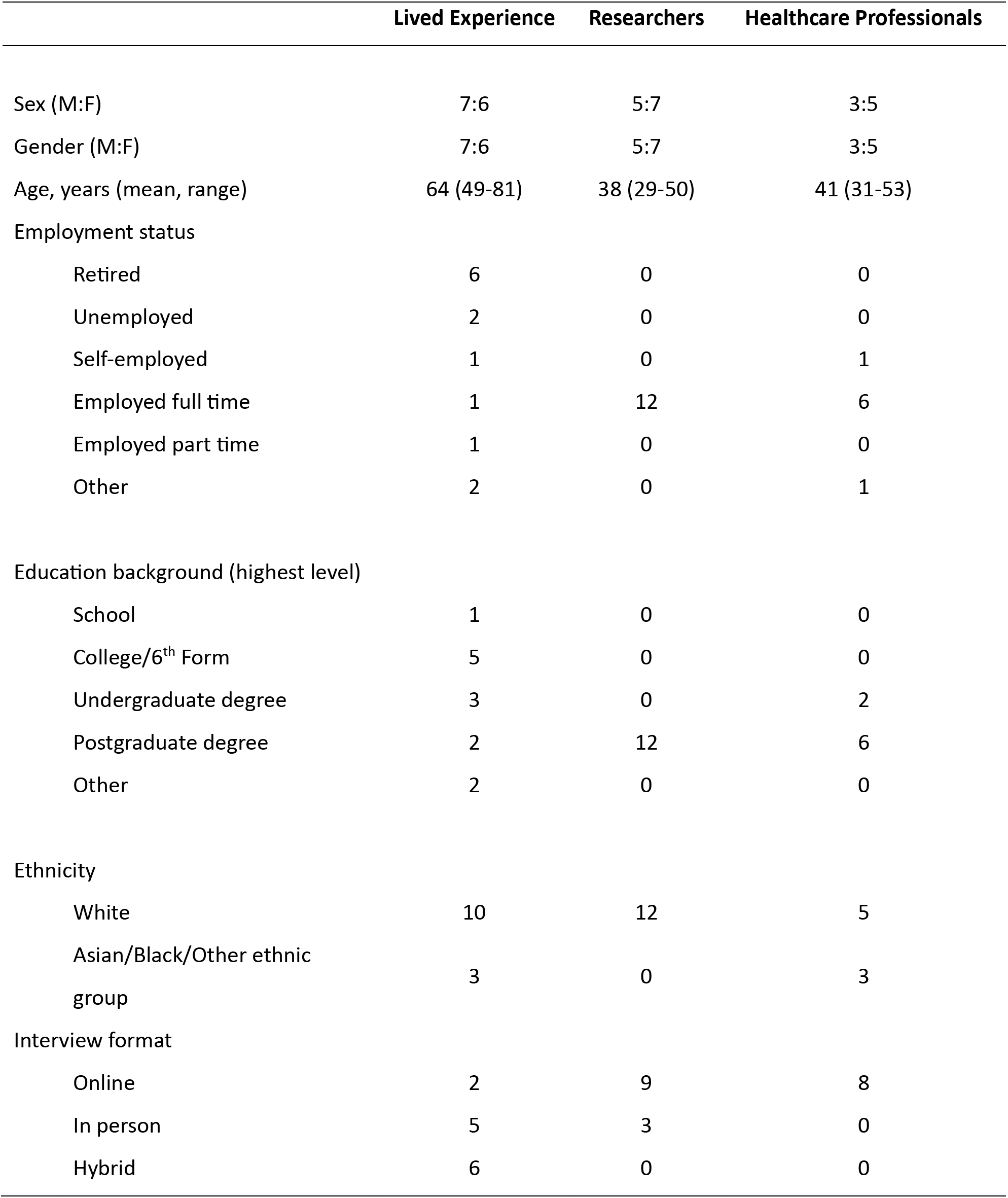
Participant characteristics.

|  | Lived Experience | Researchers | Healthcare Professionals |
| --- | --- | --- | --- |
| Sex (M:F) | 7:6 | 5:7 | 3:5 |
| Gender (M:F) | 7:6 | 5:7 | 3:5 |
| Age, years (mean, range) | 64 (49-81) | 38 (29-50) | 41 (31-53) |
| Employment status |  |  |  |
| Retired | 6 | 0 | 0 |
| Unemployed | 2 | 0 | 0 |
| Self-employed | 1 | 0 | 1 |
| Employed full time | 1 | 12 | 6 |
| Employed part time | 1 | 0 | 0 |
| Other | 2 | 0 | 1 |
| Education background (highest level) |  |  |  |
| School | 1 | 0 | 0 |
| College/6<sup>th</sup> Form | 5 | 0 | 0 |
| Undergraduate degree | 3 | 0 | 2 |
| Postgraduate degree | 2 | 12 | 6 |
| Other | 2 | 0 | 0 |
| Ethnicity |  |  |  |
| White | 10 | 12 | 5 |
| Asian/Black/Other ethnic group | 3 | 0 | 3 |
| Interview format |  |  |  |
| Online | 2 | 9 | 8 |
| In person | 5 | 3 | 0 |
| Hybrid | 6 | 0 | 0 |

Data collection took place over 5.5 months, and recruitment was stopped once we reached a point of informational power that enabled development of themes. The study was approved by the local Research Ethics Committee (Reference MSIDREC 880646) and all participants provided electronic written informed consent prior to participation.

### Procedure

Following informed consent, participants completed a demographics and experience questionnaire online (using Jisc survey software). As part of this they rated their baseline knowledge of brain stimulation technologies from 1=none at all, to 10=extensive. They were also asked to choose which brain stimulation technologies they had heard of from a list (with an additional free text option) and to describe in a few words what brain stimulation meant to them.

Participants then attended one focus group interview lasting approximately two hours. The interview was audio-recorded and transcribed verbatim by an engagement professional with assistance from the automatic transcription tool in Microsoft Teams. Participants were compensated for their time and travel (if applicable).

The interviewer (MW) is a male postdoctoral researcher with a PhD in Sensorimotor Neuroscience. Additional questions were raised by MKF, who is a female Associate Professor with a PhD in Human Neurophysiology. Both MW and MKF have worked extensively with NIBS, particularly tES and TMS, and with people with brain injury, particularly stroke. Their research interests are focused around understanding and modulating motor learning and motor recovery, as well as sleep.

The topic guide (table 2) was initially developed by MKF, with input from MW and members of the public from our research centre’s Brain Health Advisory Group. The topic guide was used to ensure coverage of important points, but the direction of the interview (and order of topics) was largely dictated by the conversations of the group members. The interview began with an explanation of the overall aims of the project (the focus groups are a part of a larger project to develop recommendations for translation) as well as the purpose of the focus group interviews, followed by a brief presentation describing the NIBS techniques of interest. Participants were encouraged to interrupt during the presentation to ask for clarifications or provide their thoughts about the points raised.

**Table 2.** Topic guide for focus group interviews.

|  |
| --- |
| 1. Understanding of NIBS technologies |
| 2. Experience of using NIBS (themselves or others they know) |
| 3. What do we need to know? |
| 4. What would make NIBS more or less useful? |
| 5. Comparison to pharmacological treatments |
| 6. Safety and efficacy |
| 7. Optimism, excitement, concerns, worries |
| 8. Language and terminology |

### Analysis

Interview data were analysed by MKF using a reflexive thematic analysis approach as outlined by Braun and Clarke ^10^. An inductive approach was used, but we acknowledge that the researchers had some pre-existing ideas based on previous studies and experience. Transcripts were not provided to participants, but a general overview of findings were presented to five participants who could provide comments and raise any questions before the final draft manuscript was prepared.

Transcription, familiarisation, and initial coding were conducted in parallel with participant recruitment. Thematic identification occurred throughout data collection. Regular debriefing between team members enabled reflection, ensured rigour, and helped guide the latter interviews (e.g. to identify concepts that needed to be explored more deeply, or to probe how perceptions might differ between groups). However, the themes were not finalised until all data collection had stopped, when MKF and MW reviewed the findings to refine the themes, ensuring that they accurately reflected patterns supported by the data.

## Findings

Demographic information can be found in table 1. In total, 33 people were interviewed across the three participant groups (LE: n=13, R: n=12, HCP: n=8). One researcher also identified as an HCP, and two HCPs were researchers. Two researchers and one HCP also reported lived experience of brain conditions outside of their professional experience.

Pre-interview self-reported ratings of understanding of NIBS and example quotes for the question “what does brain stimulation mean to you” can be found in table 3. As anticipated, familiarity of NIBS was highest for the researcher group and lowest for the LE group. Six LE participants reported not having heard of any brain stimulation technologies, three had heard of tES, and three had heard of TMS. Most HCPs had heard of TMS, and five were familiar with tES.

**Table 3:** Pre-interview ratings of perceived understanding of brain stimulation technologies from the three participant groups.

|  | Rating of NIBS<br>understanding pre-interview (out of 10) |  | What does brain stimulation mean to you? |
| --- | --- | --- | --- |
|  | Mean | Range |  |
| **Lived Experience** | 3.4 | 1-8 | *“Using some sort of electric means to try and stimulate certain parts of the brain, in order to help with particular conditions or functions of the body”*<br>*“Externally introduced stimulation may be necessary for learning but we have not always done it well and there are many individual and societal dangers so we must learn well.”*<br>*“Something that rejuvenated your brain cells”*<br>*“A way of using external stimulation to explore brain wave patterns. Use of external stimulation to correct behaviours”* |
| **Researchers** | 9.0 | 8-10 | *“A way to modulate brain activity and a great candidate for clinical intervention”*<br>*“Brain stimulation is used in research to investigate the role of specific brain areas, or in therapeutic contexts to regain healthy brain function.”*<br>*“Useful research tool; exciting set of techniques with lots of promise, hype and disappointments”* |
| **Healthcare Professionals** | 6.3 | 3-10 | *“A tool to modulate brain activity with potential to enhance recovery.”*<br>*“The use of electrical or magnetic energy/charge to alter local or global neurological function with the aim of improving mood, motor or cognitive function”*<br>*“A means of altering or influencing brain activity externally.”* |

Seven themes were developed. Example quotes and codes underlying themes are presented in Figures 1-3 and thematic findings are described.

**Fig 1.**
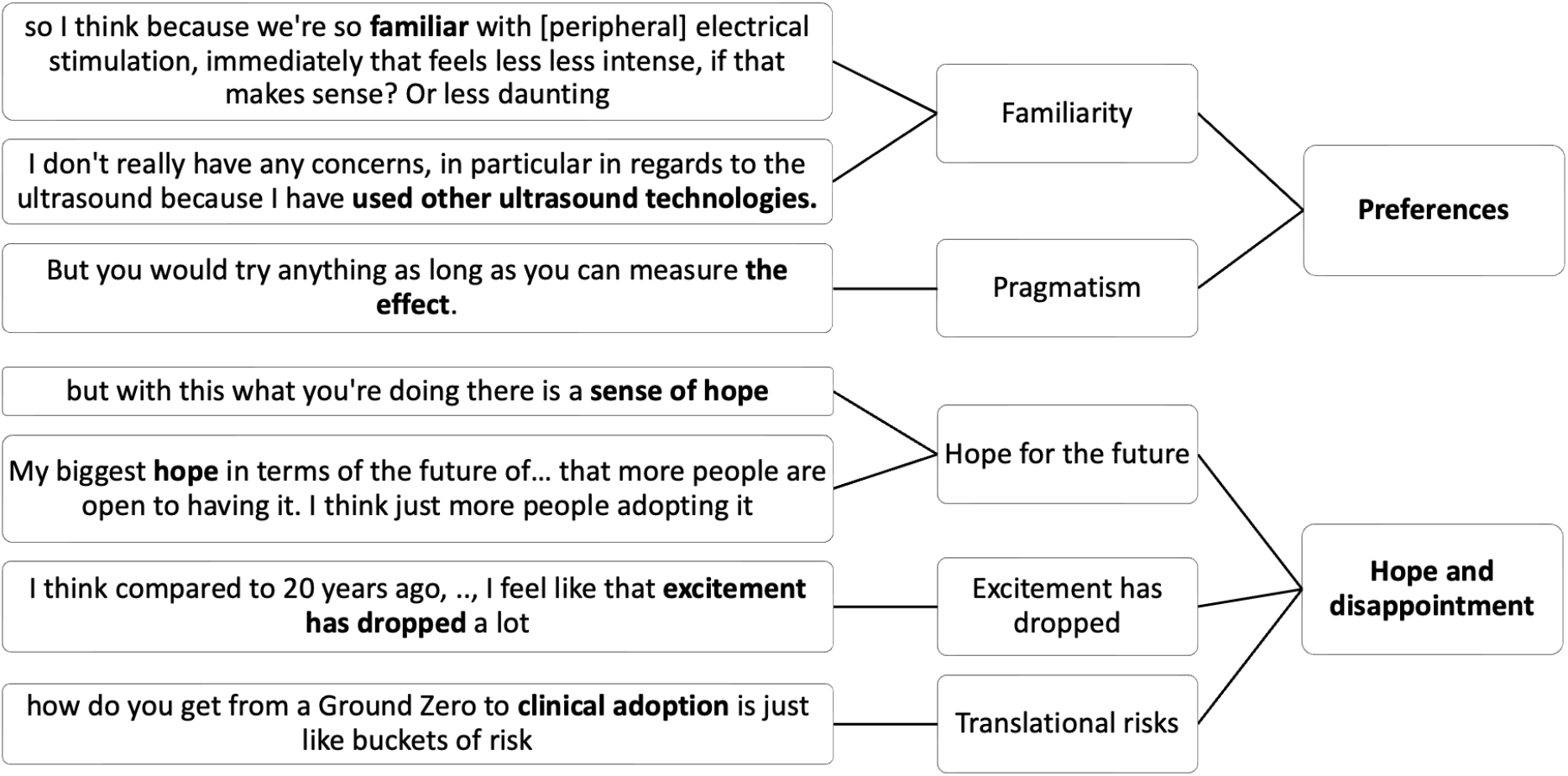
Example quotes (left) and codes (middle) for “Preferences” and “Hope and disappointment” themes.

### Preferences

Participants in the LE group felt that the non-invasive nature of the stimulation was reassuring.

> *LE: “It’s kind of the word is non-invasive, which I really like. You do it, you get up and you go, you know? So that’s reassuring as a patient.”*

However, there was a general lack of consensus on which technique was more preferred, with frequently conflicting rationale. For example, some felt TUS to be scariest because it targeted deep regions, whereas others preferred TUS for that reason.

> *LE: “[TUS], was the scariest one for me, because it went deep.”*
>
> *LE: “because it [TUS] goes deeper, you know, into the brain and in my head, …… I think maybe because it got can go deeper, you might even need less sessions.”*

Some participants also preferred a particular stimulation approach because they felt it was familiar, such as ultrasound for pregnancy or because they had used peripheral electrical stimulation.

> *LE: “I have used other ultrasound technologies. Obviously I’m a mother so, you know, it’s kind of routine that we have our scans.”*

Many expressed a pragmatic approach that the most important thing was that it worked.

> *LE: “But in in short. Whatever works. “*
>
> *R: “I would rather not say, well, let’s ditch all of this technique and focus all our efforts on the other one.”*

### Hope and disappointment

All groups discussed the hope and general promise of NIBS.

> *LE: “I would love to be able to go back to my friends and say this is what’s going on and maybe it will go into the NHS.”*
>
> *HCP: “So I really hope you guys manage to work out how to push it forward because at the moment it does seem like pharmacological things are being pushed much more than electrical devices.”*

However, researchers in particular acknowledged fluctuating excitement and disappointments resulting from slow progress. Participants highlighted persistent barriers as well as technical, regulatory and market risks associated with pursuing NIBS for clinical adoption.

> *R: “I’ve been looking at when the first TMS studies were done and they were done in the 80s, right? Like it’s 45, almost 50 years now. And I mean, we’re a lot better at understanding how it works and having different protocols than we were in the 80s, but ….”*
>
> *R: “how do you get from a Ground Zero to clinical adoption is just like buckets of risk. So, there’s a technology risk, like can you build the actual thing? You know there’s a clinical risk - does it actually work? There’s a regulatory risk - like can you get it past the regulations? And then there’s like a reimbursement risk - like can somebody pay for it? And then there’s like an adoption risk - like a market risk.”*

### Communication

All groups identified a need for improved communication, awareness and understanding for patients and the public as well as healthcare professionals. There was a perceived lack of understanding or knowledge about NIBS and participants felt this might provoke people to worry about risks. This lack of knowledge was also believed to increase the likelihood that healthcare professionals would prescribe more familiar treatment instead.

> *R: “The problem is obviously that many patients are not still familiar with the idea of brain stimulation that might be worried about the risks, et cetera.”*
>
> *LE:“.. I think I would be hesitant if I didn’t understand what it was, how it was going to work.”*
>
> *HCP: “I think for me the biggest barrier is probably awareness. If we had more people who actually knew about TMS and its benefits, and it’s not as scary as you think it is, we’d probably have more people adopting it.”*

All three groups highlighted the importance of explaining that NIBS is not directly comparable to electroconvulstive therapy (ECT), due to the fear associated with ECT – largely driven by its portrayal in popular culture and film. However, HCPs discussed that ECT is clinically very effective and should not be overlooked.

> *LE: “You cannot just say, ‘oh, we’ve got electrical things.’ I mean, go. People go, “Oh, my God. Are you crazy? This is back to the 30s or whatever it is.” It’s a big, big, big, big fear, a big fear. Huge fear.”*

Researchers acknowledged that there was a balance to strike - the more complex the technology, the harder it would be to communicate clearly. Terminology may need to be different for precise stimulation protocols in comparison with low-level repeated protocols administered at home. Healthcare professionals who were already using NIBS did not feel that explaining the technique was a major barrier.

> *R: “I think we should be honest with what it does and what we do and do not understand about it, but it’s finding the balance between simplicity and you know, a patient will actually read the leaflet and actually want to know versus overwhelming them with information.”*
>
> *R: “If it’s a sticker, if it’s a disposable, you know supplement, then you would use a very different language I guess. If it’s a treatment, possibly over weeks, delivered by an expert, then again that that is very different.”*

All groups suggested positive stories and real-life examples could support communication.

> *LE: “That will help me in terms it will break down fear, the safety issues that maybe perhaps patients may have. You know, they see some videos, some somebody has used it and they can see, you know, not necessarily the effect of it, but just that another human being has actually used it.”*
>
> *HCP: “But if you can do that and capture it with maybe videos of patients, success over treatments and put that up in a seminar or interview or show that and demonstrate it, and if there is a patient, for example, who in live time is getting a benefit from it, use it. To show them that and demonstrate that. Then you’ll get that buy in.”*

They also discussed creating leaflets to communicate risks of side effects, like those provided with medications.

> *R: “what came to my mind now was the little leaflet you get with every drug, you know, and the sheer amount of things it covers for.”*

Both researchers and HCPs highlighted the need to improve knowledge for clinical teams, policy makers and commissioners. They specified that an intervention needed to be clearly communicated in guidelines to be usable, with a directive from ‘people at the top’.

> *R “Because I’m, I’m worried if doctors don’t know about it then policymakers won’t know about it either.”*
>
> *HCP:”.. we always say go back to the evidence pull on the evidence. Is it sufficiently powered? Is there a Cochrane review to support this intervention? Is there a guideline that you can refer to?” HCP: “In a way to start anything new in the NHS, you sort of need top down pressure, whether that’s from NHS England or…”*

Training was seen to be needed but not necessarily perceived as a barrier to translation by researchers or HCPs.

> *HCP: “I don’t think in terms of delivery would be particularly difficult to train people up.”*

### Accessibility

Cost was identified as a key barrier to overcome for NIBS to be accessible, though researchers tended to feel that the issue was that cost-effectiveness was yet to be robustly demonstrated.

> *LE: “you’re still all trying to think what might be best or what might be most efficient. Being honest, it’s the NHS, so what might be more cost efficient as well?”*
>
> *R: “I think no one has really sort of done the calculations in terms of, you know cost economics.” HCP: “show now as well with a tighter budget that it’s going to either shorten the patients length of stay, improve their outcomes, reduce the carer burden, so it really demonstrate the cost effectiveness of the intervention.”*

Researchers highlighted that whilst accessibility barriers exist, there was promise for the future. tES was flagged as leading the way for home use, along with real potential for “helmet-type” devices for tUS on the horizon. LE participants identified that for stimulation to be used in the home, it would need to be easy.

> *R: “that was always the big advantage of electrical stimulation is it can be put in the home fairly simply.”*
>
> *R: “the ultrasound… there’s a lot of companies springing up that are wanting to ultimately build wearables, right.”*
>
> *LE: “But if you can show if you if you can show well, ‘this is as easy as it is like putting your glasses on. Not a big thing, you know.”*

It was acknowledged that there needs to be considerations with regards to issues of compliance for home-based NIBS, and all participant groups felt that there were potential advantages to being delivered in a clinical setting.

> *R: “So people say ‘I want low cost. I want wearable. I want non-invasive.’, but consumer behaviour or like medical consumer behaviour doesn’t match those sentiments, right?”*
>
> *LE: “And in terms of having it at home, I don’t trust myself to use it. You know, I’ll be distracted and I might get talked out of it.”*
>
> *LE: “sometimes having a trusted, you know, someone that the patient sees it’s a trusted professional administering treatment can be an advantage, and then that would take some of the responsibility away from the carer as well.”*

Researchers and HCPs also identified that there are technical barriers to overcome to ensure accurate treatment in a home setting and felt it would be important to continue to pursue both precision and scalability.

> *R: “if these things do translate into a home environment, how do you make sure that people are applying it properly? So I think that’s probably a really big technical challenge of, like, you’ve got to, if it’s so focal, you’ve got to get it in the right place.”*
>
> *R: “You might end up having things at two different ends. You have neuromodulation of the kind we’re talking for, very specific applications where you may need precision … and then there’s the the other extreme of the spectrum which say is is a sort of sticker based disposable stuff in in it’s very extreme form, that can be deployed on a mass scale and anybody can stick it on for anything if they they want to…”*

**Fig 2.**
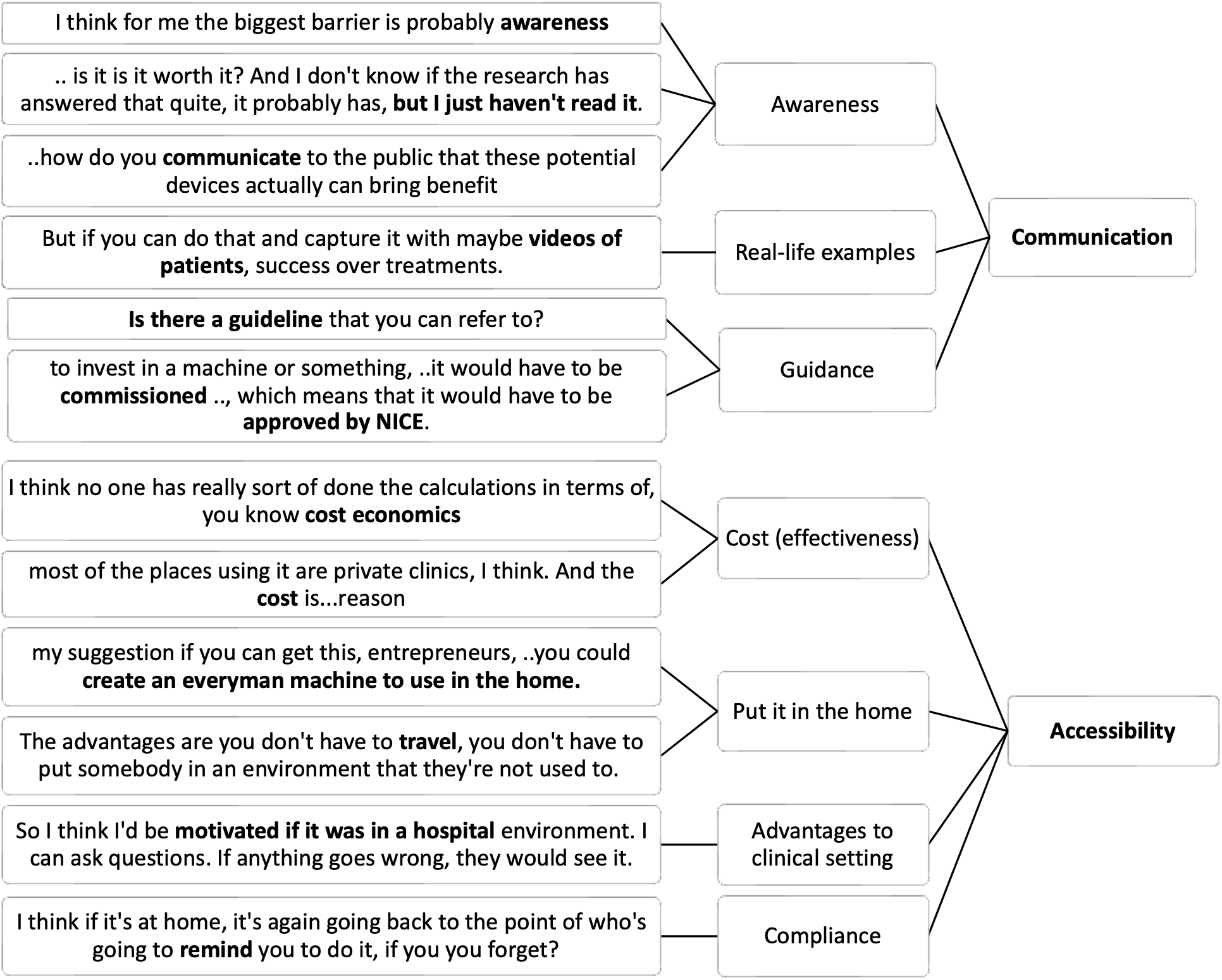
Example quotes (left) and codes (middle) for “Communication” and “Accessibility” themes.

### Infrastructure

Researchers and HCPs discussed the need for increased collaboration to push forward evidence gathering, optimisation and translation.

> *R: “But you need to suddenly be working across engineering, across neuroscience, across clinicians.”*
>
> *R: “that if you have the industry partner, then you can show a, you know, an immediate ready route to commercialization… And the ability to, you know, to scale and roll things out.”*

However, collaboration was seen to be difficult, particularly between industry and academic partners.

> *R: “Yeah, I think that that’s come up with various kind of industry-academic collaboration things… like the collaborations themselves but also the just the discussion of how we can make that better in the university and often these things yeah the IP and who owns it is can be a stumbling block.”*
>
> *HCP: “think advocating for an an easier way to collaborate and share data between companies and researchers, I think would make make it much easier to actually produce the evidence base that is needed for a clinical device to be marketed.”*
>
> *R: “… but typically what you want is speed and universities are not well known for speed.”*

There was also a perception that the structure of academic research poses challenges for sustained progress and longitudinal evidence of effects, with short-term roles and limited continuity making long-term, large-scale studies difficult to deliver.

> *R: “So it’s it’s also like getting enthusiasm and excitement within the group to run this kind of study, which is more like a slow-moving snail rather than the fast-moving horse that we all want in science.”*
>
> *R: “think needing big funding needing the long timespan, which often we don’t even have in our contracts, right. At least if you’re at a more junior level. So the - a longer term support from both structurally, financially and people in the group. I think that’s maybe the main stumbling block.”*

Finally, HCPs discussed the anticipated difficulties with NHS infrastructure. It was felt that setting up a service to deliver NIBS would be difficult, and delivery would need to span multiple conditions to be achievable.

> *HCP: “like starting a new service, in my experience has been quite difficult.”*
>
> *HCP: “I think it might, it probably might, it would be much easier to have a service that deals with all forms of stimulation, like having a stimulation suite in a sense in a hospital rather than having TMS in the psychiatric clinic or and ultrasound in the neurology clinic and whatever in another clinic, I think you would kind of need to have that expertise shared across different modalities.”*
>
> *HCP: “you’re going to have to sell this not just as a treatment to be used for mental health patients, but also across pain and neurology.”*

### Ethical and regulatory uncertainty

Generally, researchers and HCPs didn’t feel that NIBS raised unusually high ethical barriers, relative to other techniques. They highlighted the necessity to provide clear materials to facilitate decision making and also acknowledged that patients who are ‘desperate’ might be willing to try anything.

> *R: “But I don’t think that there’s anything special about neurotechnology. I think that there’s all sorts of devices out there that collect way more sensitive data and information about people than anything we’re going to be able to do…”*
>
> *HCP: “I think that’s probably an issue with neurodegenerative disease, like people are quite like, the patients are quite desperate. So they’re quite keen to try anything that is offered to them.” LE: “But you would try anything as long as you can measure the effect.”*

However, it was acknowledged that there are likely to be unforeseen ethical issues arising in the future as NIBS develops and changes, which should not be overlooked.

> *R: “And the kind of unique ethical issues that we might like to chew over are the things that are not quite here, but which we can reasonably predict in the medium future.”*
>
> *R: “but it’s interesting that these types of question about what is important, what is valued and what should be protected have come up when we have seen the emergence of specific neuro-technologies that might have direct effect.”*

Additionally, some researchers, and HCPs, acknowledged a wariness when it comes to medical device regulations, which were felt to be less clear than medications and better understood by industry partners.

> *R: “It’s [regulatory approvals] a barrier if you’re within an academic institution. So if you had to do an MHRA device submission, like for a clinical investigation of a medical device and you would just built something in the lab, it’s going to be extremely difficult for you to compile the dossier of information that you need to get that approval, without at least a lot of effort and a lot of, you know, external consultancy… So I think for a company it’s a different viewpoint. So from our side we don’t see it as a particular barrier, like we have to do this stuff anyway.”*
>
> *HCP: “Yeah, I find the whole medical devices field is a little bit less clear than medications.”*

### Evidence base

All groups discussed the balance between knowing how NIBS works, *vs* knowing that it works. Many described a need to understand the mechanisms primarily for optimising protocols.

Ultimately, knowing ‘that it works’ and the associated longitudinal changes, is more important to the people receiving it than understanding the precise neurophysiological effects of the stimulation. It was felt that the mechanistic research should continue in parallel and shouldn’t further delay translation of effective techniques.

> *LE: “the priority has to be to, we are looking to see if it works and what way it works, but for how long it works, who it works for.”*
>
> *R: “I don’t think these things are going to be like just a one off treatment strategies like with many other things like talking therapy or taking pills, they will they need cumulative, and they need. Yeah, they need a longer period of testing and application”*
>
> *R: “I’d say there’s a balance between exploring the parameter space, but also not waiting for that to happen before we take something, even if it’s a local optimum and not the global, into a clinical translation, because otherwise we will never get there.”*
>
> *HCP: “I think it’s one of those things that it would be nice to know, but actually the efficacy is proven and the safety is proven and those are more relevant, I guess.”*

An understanding of how NIBS works, and in particular the potential unintended effects the stimulation could induce, was also seen to be important to ensure safety of delivery and informed consent.

> *R: “I don’t understand how they could have consented the patients when they didn’t understand what it was that they were doing.”*

Researchers frequently discussed the challenge of parameter optimisation at the individual and group level.

> *R: “we’re talking about things that we think are going to be a gradual buildup of therapeutic effect over multiple sessions while sitting in a sort of quite a high dimensional sort of parameter space. How do you go about kind of optimising it?”*
>
> *R: “I mean, there’s a lot of different aspects that you can optimise on, right.”*
>
> *R: “in a way it kind of almost started with everyone using the same protocol and now people realising maybe the same protocol doesn’t actually work for everyone.”*

**Fig 3.**
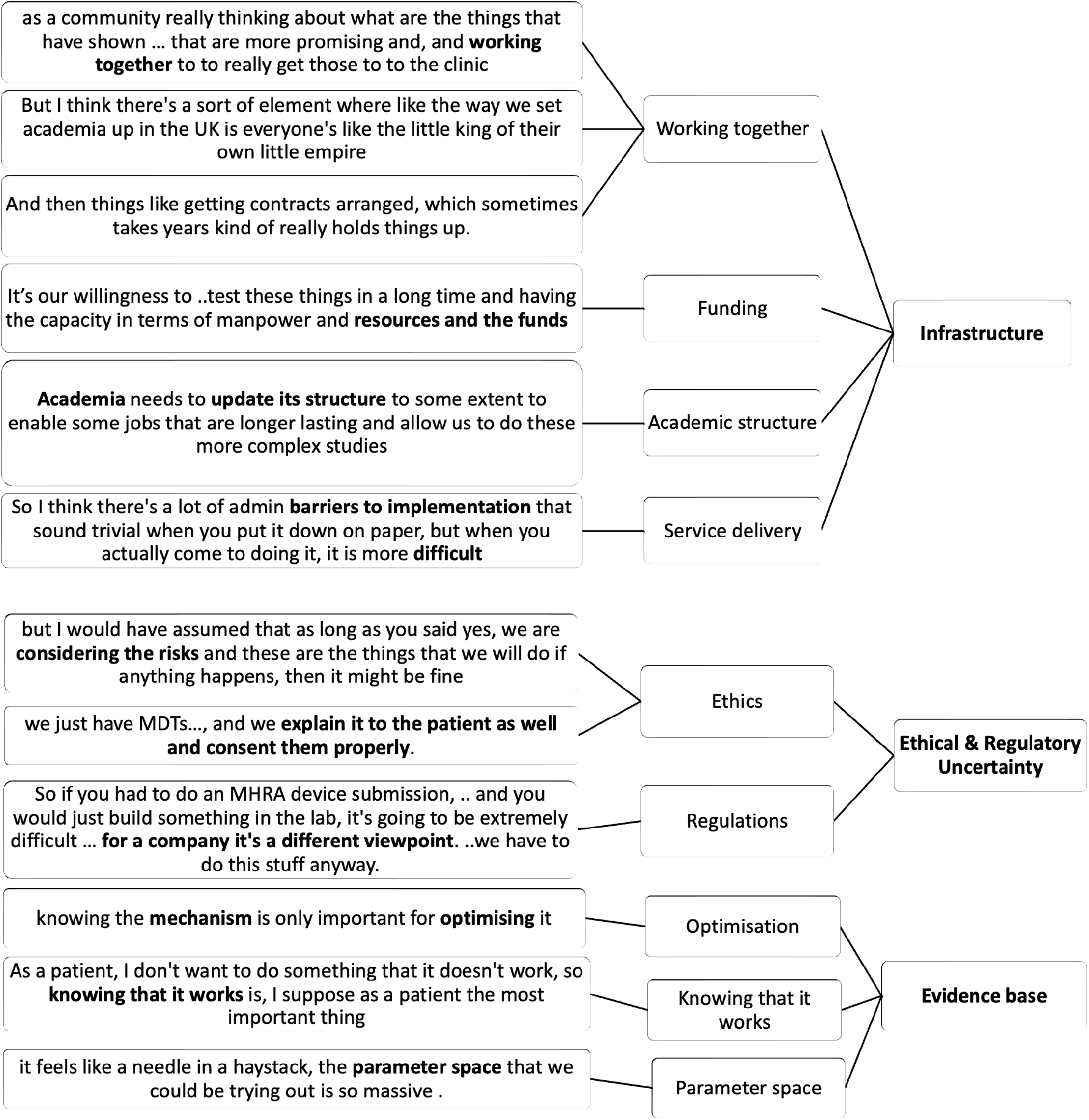
Example quotes (left) and codes (middle) for “Infrastructure”, “Ethical & Regulatory uncertainty” and “Evidence base” themes.

## Discussion

We utilised focus group interviews to explore perceptions of non-invasive brain stimulation and potential barriers to clinical translation across neurological, neuropsychiatric and neurodegenerative conditions. We included people with lived experience of three key brain conditions (brain injury, depression and dementia), as well as researchers and healthcare professionals working in these areas to obtain a range of opinions. Across stakeholder groups, several cross-cutting themes emerged, including positive perceptions towards NIBS and future opportunities, as well as a range of perceived barriers.

There was a general lack of consensus in terms of preferences for stimulation approaches, and where preferences did exist, they tended to be driven by factors such as perceived familiarity, established use-cases (e.g. rTMS already being approved for depression), and depth of action (with “depth” being inconsistently associated with a preference or concern). This is in contrast to Atkinson-Clement^9^ who found that participants tended to hold a stronger preference for tUS. However, the basis for their observed preference seems to centre, at least partly, on a belief that ultrasound is most effective and safest, which was not consistently found here. We therefore propose their findings may reflect participants’ familiarity with ultrasound for pregnancy, leading them to equate tUS with safety. Many participants in the current study ultimately endorsed a pragmatic “whatever works” stance, highlighting that it is important to continue to pursue all avenues in order to establish the option that works best for a particular condition or symptom. Participants also expressed hope regarding the potential for NIBS in the future, though researchers noted a fluctuation in excitement due to persistent technological, regulatory, and market risks that are yet to be fully addressed.

A key theme was the need for improved communication with patients, the public, and within healthcare systems. In line with the lived experience preferences being heavily influenced by familiarity, improving the awareness of the general public with NIBS techniques is vital for future successful clinical translation. Participants emphasised that low public awareness may provoke people to worry about risks. This is consistent with Cabrera et al ^6^ who identified a lack of knowledge and awareness to be a barrier to neurostimulation more broadly, and Atkinson-Clement ^9^, who found that providing brief information increased ratings of optimism and comfort, while reducing confusion. The NIBS community may benefit from learning from the experience of ECT, where years of negative media and public portrayals have contributed to misconceptions and fear among patients and the public ^11^. This variability in awareness was also reflected among healthcare professionals included in the study, who reported a vast range of pre-interview experience with NIBS, from extensive familiarity to very limited exposure. We speculate that this potentially reflects academic involvement with those involved in research more likely to be exposed to newer techniques, either directly or through conference attendance. This disparity in awareness further stresses the need for clear guidelines, top-down endorsement, and clinician education to support the prescription of NIBS, when appropriate, over other more familiar treatment options.

Accessibility barriers centred on cost effectiveness, user compliance, and technical hurdles to ensure accurate treatment, particularly in a home setting. While tES was seen as “leading the way” for home use, concerns remained about ease of use and how to ensure correct application. Crucially the clinical setting was still seen to hold value, particularly to provide reassurance and reduce burden on patients and carers. However, the time required to attend multiple sessions of stimulation was seen as a possible barrier for some, as has been reported previously ^6^. There were also clinical infrastructure barriers that were felt difficult to overcome. Although training of staff was not seen to be a particular challenge, the hurdles to setting up a stimulation service in the NHS were considerable, and it was difficult to see how this would be achieved without compelling evidence of cost-effectiveness. However, we argue that demonstration of cost-effectiveness is not enough.

Indeed, in the case of TMS for treatment resistant depression, there is considerable evidence of cost effectiveness, and approval for use by the National Institute for Health and Care Excellence (NICE) in the UK – yet uptake remains extremely limited ^12^.

## Limitations

Participants were limited to those with experience of research and healthcare in a UK setting. Whilst this was deliberate to understand potential barriers to implementation in the UK National Health Service, we cannot assume that these findings will directly translate to other countries with different health services. There also may be additional and different barriers to translation in low-resource settings, such as low-middle income countries.

Similarly, we chose to focus on three main conditions of interest: depression, dementia and brain injury, given their high prevalence. Whilst this approach enabled a variety of opinions, it did not provide sufficient data to identify potential condition-specific nuances, and the study was not designed to allow direct comparison between stakeholder groupings.

## Summary

This study contributes to a growing body of literature seeking to explain and understand the translational gap between decades of NIBS research and its slow uptake in a clinical setting. We highlight a range of barriers and potential solutions to address for the future.

## Abbreviations

LE: Lived Experience
HCP: Healthcare Professionals
R: Researchers

## Rights Retention

This research is funded in whole, or in part, by the Wellcome Trust [222446/Z/21/Z, 203139/Z/16/Z and 203139/A/16/Z, 224430/Z/21/Z]. For the purpose of open access, the author has applied a CCBY public copyright license to any Author Accepted Manuscript version arising from this submission.

## Author Contributions

Conceptualization: MW, JW, EM, CJS, MKF

Methodology: MW, MKF

Investigation: MW, BR, HS, MKF

Formal Analysis: MKF

Data Curation: BR, HS

Drafting the initial article: MW, MKF

Writing – Review & Editing: MW, BR, HS, AP, EM, JW, CJS, MKF

Funding Acquisition: MW, JW, EM, CJS, MKF

## Data availability

Data are available upon reasonable request to MW or MKF.

